# Chronic pruriuts on non-lesional skin does not affect the epidermal barrier – results from the SOMA.PRU study

**DOI:** 10.1101/2025.08.21.25334158

**Authors:** Evgeniia Komarova, Christian Meß, Finn Abeck, Inga Hansen-Abeck, Ewa Wladykowski, Volker Huck, Konstantin Agelopoulos, Sonja Ständer, Christian Gorzelanny, Stefan W. Schneider

**Affiliations:** Department of Dermatology and Venereology, University Medical Center Hamburg-Eppendorf, Hamburg, Germany; Section for Pruritus Medicine, Department of Dermatology, and Center for Chronic Pruritus, University Hospital Münster, Münster, Germany

## Abstract

Chronic pruritus on non-lesional skin (CPNL) is a hallmark of chronic pruritus of undetermined origin (CPUO) and characterized by persistent pruritus without visible skin lesions. While atopic dermatitis (AD) is well known to involve epidermal barrier disruption, the pathophysiology of CPNL remains poorly understood. Our goal was to compare stratum corneum (SC) morphology and its role as a functional epidermal barrier, the severity of the symptoms, the relationship between persistent itch and primary skin and scratch lesions, and selected inflammatory markers in patients with CPNL, AD, and healthy controls. We assessed corneocyte morphology and corneodesmosome density in skin samples using atomic force microscopy (AFM) and fluorescence microscopy (FLM). Transepidermal water loss (TEWL) and tissue hemoglobin index (THI) were used to evaluate epidermal barrier integrity and skin blood perfusion. Symptom severity was assessed using the Worst and Average Itch-Numerical Rating Scale (NRS), Scratch Sign Score (SSS), and patient-reported sleep disturbance. AD patients demonstrated structural differences in the SC, including reduced corneocyte area, clustering of corneocytes, absence of intermediate filaments, and relocated corneodesmosomes, along with elevated TEWL, THI values and IgE serum levels. In contrast, patients with CPNL displayed corneocyte morphology and skin barrier parameters like healthy controls, despite reporting high itch intensity and frequent sleep disturbance in clinical interviews. These findings indicate that CPNL is not triggered by a disrupted epidermal barrier but may represent different mechanisms.

## Introduction

Chronic pruritus on non-lesional skin (CPNL) is a highly burdensome condition characterized by persistent itch without visible lesions. It may represent a clinical manifestation of an underlying neurological, psychiatric, malignant, or systemic disorder, each of which requires independent diagnostic and treatment and therefore should not be disregarded.^1^ Although the molecular mechanisms promoting CPNL are poorly understood, previous studies have shown an association to include asteatotic eczema, porphyrias, dermatitis herpetiformis, Duhring, polycythemia vera, systemic mastocytosis, solid tumors (e.g., lung and breast cancer), diabetes mellitus, thyroid dysfunction (hypo-/hyperthyroidism), multiple sclerosis, celiac disease, and others.^2^ In approximately 13 to 50% of the CPNL patients the underlying disease remains unknown despite extensive diagnostic efforts, depending on the studied population.^2^ Accordingly, CPNL is most prevalent in chronic pruritus of multifactorial and unknown origin (CPUO) and comprise around 30% of the whole chronic pruritus cohort.^3^

Atopic dermatitis (AD) is a chronic inflammatory skin disease characterized by repeated skin lesions and itching.^4^ The clinical characteristics of atopic dermatitis vary depending on ethnic, age, geographical location and severity of the disease. Acute lesions may appear as circumscribed patches of eczema, characterized by papulovesicles, edema, crusting, papules, and scaling. After healing, the lesions may leave behind areas of hypopigmentation or hyperpigmentation.^5^

Although the exact mechanisms of the disease pathogenesis remain unclear, immune dysregulation and epidermal barrier dysfunction are considered to be critical for the development of AD.^6^ To which extent a disrupted epidermal barrier function promote the development of CPNL is unknown. The aim of the current study was to explore the differences in the morphological characteristics of the SC of CPNL patients in comparison to patients with AD and healthy controls. To assess morphological features of the skin, we employed atomic force microscopy (AFM) providing 3-dimensional, high resolution topographic images of the SC.^7^ We validated the AFM images by immunofluorescence microscopy-based detection of corneodesmosin (CDSN), a structural protein within the SC and essential for the physical skin barrier. Impaired CDSN function is not only associated with severe barrier defects but also pruritus suggesting a mechanistic link between itch sensation and the skin barrier.^8^ A reduced skin barrier is also considered to be a driver of AD and previous studies have linked mutations in the structural protein filaggrin to disease severity.^9^ AD severity is often connected to stress and psychological disorders such as depression or anxiety.^10-12^ Mental health might also contribute to CPNL, therefore, to link morphological changes of the SC, pruritus and mental health, we performed clinical interviews and determined the Scratch Sign Score (SSS) and Numerical Rating Scale (NRS) for pruritus.

We found that patients with AD demonstrated significant morphological alterations of the SC characterized by reduced corneocyte size, diminished numbers of intermediate filaments and relocation of corneodesmosomes. These morphological alterations correlated with an increased TEWL and tissue hemoglobin index. NRS and SSS evaluations showed significant burden of distress together with primary skin lesions and scratch lesions. In patients with CPNL, morphological parameters of corneocytes functionality of the epidermal barrier proved to be very comparable to healthy skin. Nevertheless, clinical interviews revealed that these patients experienced severe itch and sleep disturbance.

## Materials and Methods

### Study cohort

This prospective cohort study included patients with diagnosed chronic pruritus on non-lesional skin (CPNL), acute or chronic atopic dermatitis (aAD or cAD) and healthy controls. We divided our cohort into three groups. The first group included six patients diagnosed with CPNL, the second group consisted of ten patients with either aAD or cAD, and the third group comprised ten healthy controls. We compared affected (patients only) and unaffected skin of the three groups in terms of corneocyte morphology, hemoglobin levels, the mean corneodesmosome density, the mean corneocyte area and skin barrier integrity. We also assessed pruritus intensity and signs of scratching behavior. In order to define and evaluate possible intrinsic causes of itch and/or a body reaction to external factors in the pathogenesis of chronic pruritus, we measured the concentrations of IgE in blood.

The study was approved by the Ethics Committees of the Medical Associations Hamburg (2020-10200-BO-ff) and Westphalia-Lippe/Westphalian Wilhelms University Münster (2020-676 f-S), Germany. The study will be conducted in accordance with the WMA Declaration of Helsinki, guidelines for Good Clinical Practice, national and local laws. All participants are required to provide written informed consent.

### Scratch Sign Score (SSS)

In order to explore pruritus severity, we implemented the SSS, a clinical scoring method used to assess scratch-related skin lesions. It evaluates two main components with points ranging from 0 to 4 and 0 to 5 respectively. In the first part of the evaluation the primary morphological lesion type is assessed, with 0 points being granted when no lesions can be found and 4 points defining excoriated papules, nodules and/or plaques. The second component is a measurement of the body surface area affected by scratch lesions, which is estimated using the hand surface method, where 1 hand equals approximately 1% of the body surface area. 0 points are given out when patients have no scratch lesions and 5 points show that 80 to 100% of the body surface is affected. In the scoring system, the two individual scores are multiplied to define a total score.

### Numerical Rating Scale (NRS)

To assess the worst and average itch experienced during the previous 24 hours, we implemented the NRS as a self-reported item. The NRS based on the question: “On a scale from 0 to 10, where 0 indicates “no itch” and 10 indicates “worst itch imaginable”, how would you rate your itch at its worst in the last 24 hours?”^13^ An analogous question was asked to define an average itch in the past 24 hours. A clinical interview about the effects of itch on the quality of sleep was conducted.

### Tape stripping

To compare corneocyte morphology in the SC, we used tape stripping to collect corneocyte samples.^14^ Skin areas of interest on the central flexor forearm were rinsed with warm water and gently dried using soft tissue paper. A tape was then evenly pressed onto the skin and thin layers of the SC were obtained by peeling off the tape strips (Crystal Clear, Tesa SE, Hamburg, Germany). The procedure was repeated 10 times (tape strip 1 – 10) on the same site of the skin, as published previously.^15^ Tape strips 8 to 10 were analyzed by AFM and FLM.

### Atomic force microscopy (AFM)

Tape strips were mounted on a glass object slide by double-sided tape and imaging with the AFM (NanoWizard, JPK instruments, Berlin, Germany) was performed without further preparation and as previously reported^7^. The scanning probe (CSC37, MicroMasch, Innovative Solutions, Sofia, Bulgaria) had a nominal spring constant of 0.3 N/m. The skin samples were imaged in contact modus in air with a line rate of 1 Hz. AFM images were analyzed with JPK image processing software (version 3.3.20; JPK instruments AG, Berlin, Germany), gwyddion (version 2.14, free software supported by the Czech Metrology Institute) and with imageJ software, version 1.54p.

### Fluorescence microscopy (FLM)

After AFM, tape strips were further processed by indirect immunofluorescence staining using a primary polyclonal rabbit antibody directed against human corneodesmosin (CDSN, ThemoFisher scientific Waltham, USA) and a secondary Alexa Fluor 555-conjugated goat anti rabbit antibody (ThermoFisher scientific). After staining and prior to microscopy, tape strips were washed with ultra-pure water and air dried. Dry tape strips were images using an FLM (Observer z.1, Zeiss, Oberkochen, Germany) and analyzed with imageJ software, version 1.54p.

### Trans-epidermal water loss (TEWL)

Skin barrier integrity was assessed by measuring TEWL, which represents the amount of water that evaporates from the SC per area of skin. ^16^ We placed the TEWL probe (Tewameter TM 300; COURAGE + KHAZAKA electronic GmbH, Cologne, Germany) on the skin surface and measured water vapor density over a defined skin area within 30 seconds under ambient room humidity conditions, as previously described^17^.

### Hyperspectral imaging (HSI)

Hemoglobin levels in the skin were measured by HSI (TIVITA Tissue; Diaspective Vision GmbH, Am Salzhaff-Pepelow, Germany). Measurements were taken from a defined skin area (20mm) on the right or left inner forearm or hand. Image data were analyzed with the TIVITA Tissue Suite software.

### Statistical analysis

To assess significant differences between the two groups, an ordinary one-way ANOVA with Tukey post hoc test, Kruskal-Wallis test or Pearson correlation tests were conducted. A *P* value of less than 0.05 was considered statistically significant.

## Results

Compared to healthy controls (NRS = 0) both patient groups AD and CPNL showed mild (NRS > 0; NRS < 4) and high (NRS ≥ 7) NRS levels (**Figure 1A**). In contrast to the NRS, the scratch sign score (SSS) representing the strength of scratch signs at the skin, was absent in most patients and only low (SSS < 5) in 30% of the CPNL patients and comparable to the healthy control group (**Figure 1B**). In the AD group, SSS was above five in most of the patients indicating significant scratching. Despite the absence of scratch signs, itch burden was significant in CPNL patients as indicated by a strong correlation between the sleeplessness index and the NRS (**Figure 1C**).

**Figure 1:**
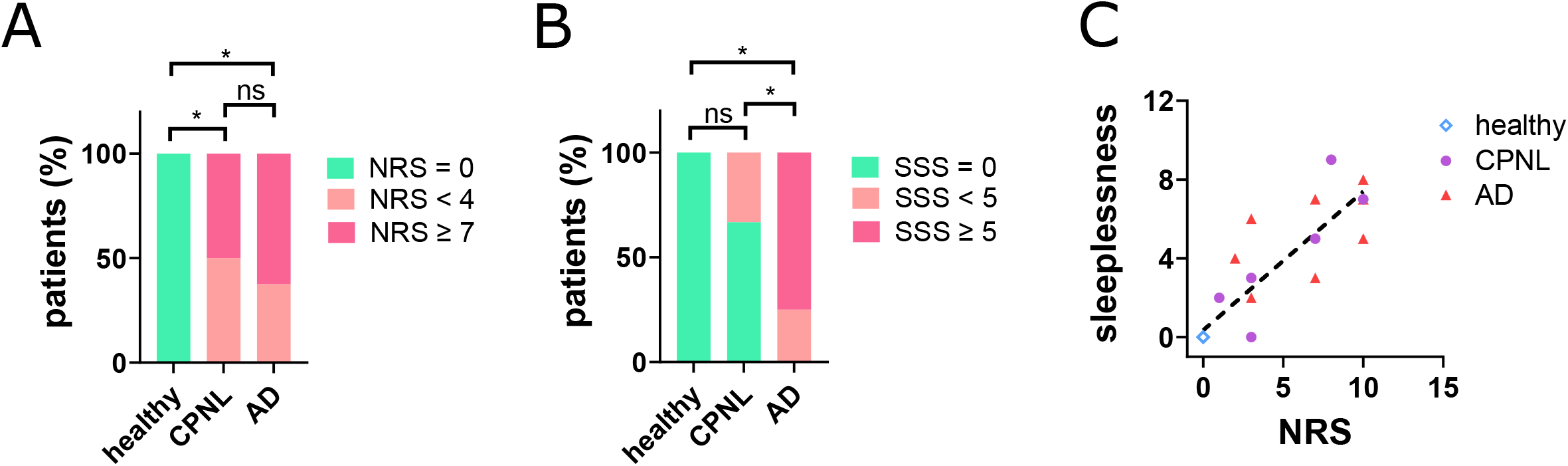
CPNL and AD patients show distinct scratching behaviors. (**A**) Compared to healthy controls (n = 10), CPNL (n = 6) and AD (n = 10) patients showed a similar NRS. Individuals were divided into three groups: NRS = 0, NRS < 4 and NRS ≥ 7. ^*^ *P* < 0.05, Fisher exact test with Freeman-Halton extension, ns = not significant. (**B**) Indicated by the SSS, patients suffering from CPNL (n = 6) showed significantly less scratch signs than AD patients (n = 10). Healthy controls (n = 10) showed no elevated SSS. Individuals were divided into three groups: SSS = 0, SSS < 5 and SSS ≥ 5. ^*^ *P* < 0.05, Fisher exact test with Freeman-Halton extension, ns = not significant. (**C**) Correlation between the sleeplessness index and the NRS showed a significant positive correlation. *P* < 0.001, two-tailed Pearson correlation test. (**D**) HSI indicate that skin inflammation was only evident in lesional regions of AD patients. In CPNL patients, pruritic (CPNL_pruritic_) and non-pruritic (CPNL_non-pruritic_) skin regions were distinguished. In AD patients, lesional (AD_lesional_) and non-lesional (AD_non-lesional_) skin regions were distinguished. ^*^ *P* < 0.05, ^**^ *P*< 0.01, one-way ANOVA with Tukey post-hoc test.

Consistent with previous literature^18^, we found elevated IgE levels in the serum of AD patients (**Figure 2A**). In CPNL patients, IgE levels were not increased compared to healthy controls. Using hyperspectral imaging (HSI), we quantified the amount of hemoglobin as a marker for blood perfusion in the skin of AD and CPNL patients and healthy controls (**Figure 2B**). We distinguished different skin areas according to disease activity. We found significantly elevates blood perfusion in the lesional skin of AD patients (AD_lesional_), indicating the inflammatory response of the disease affected skin region. Compared to healthy controls, inflammation was neither detected in non-lesional skin of AD patients (AD_non-lesional_), in skin regions of CPNL patients with itch (CPNL_pruritic_) nor in regions without itch (CPNL_non-pruritic_).

**Figure 2:**
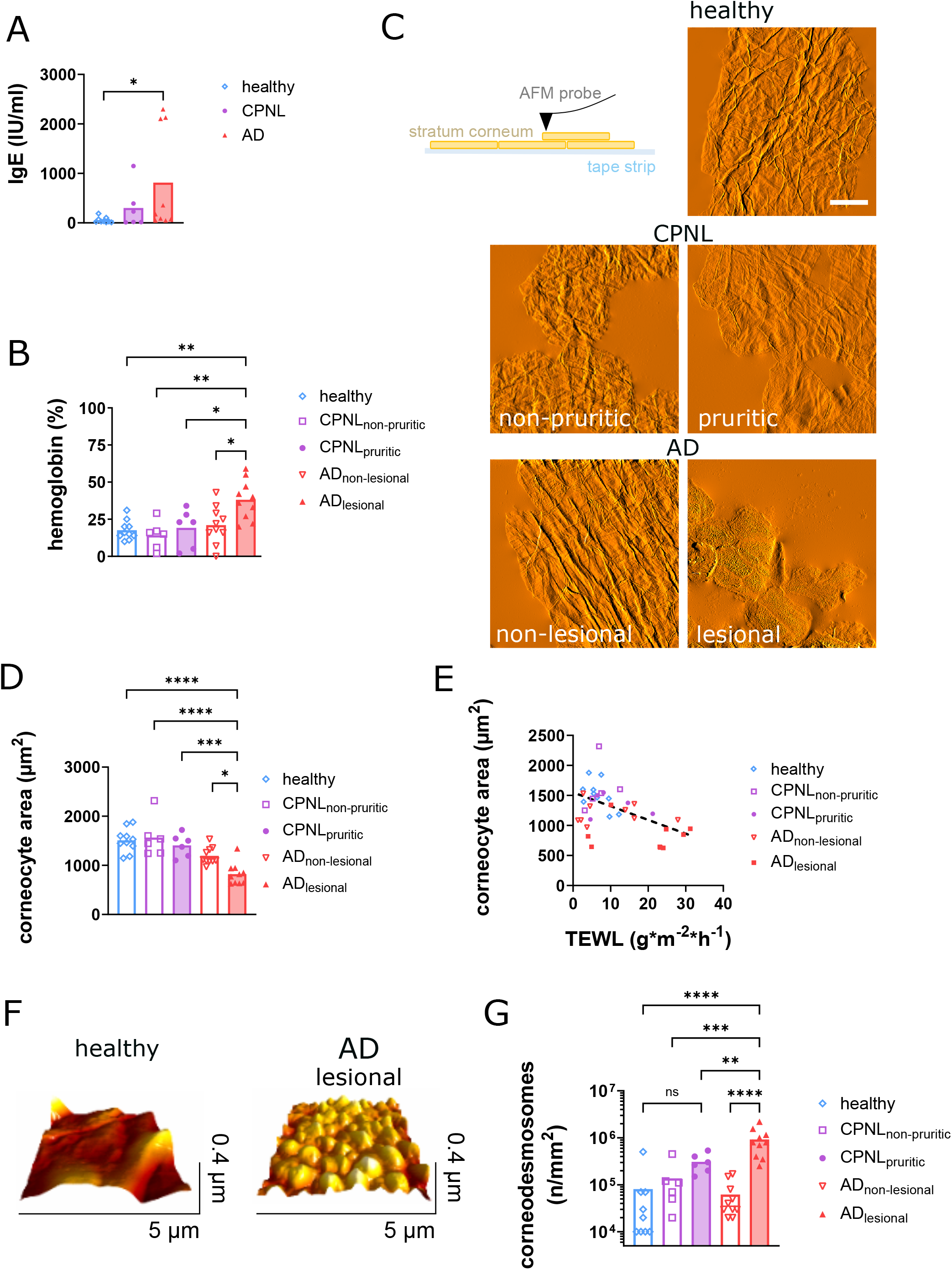
AFM imaging of the SC of CPNL patients showed no morphological alterations. (**A**) Serum IgE levels in healthy controls and CPNL and AD patients. ^*^ P< 0.05, Kruskal-Wallis test. (**B**) Skin hemoglobin levels measured by HIS in healthy controls, patients suffering from CPNL or AD. ^**^ P< 0.01, ^***^ P< 0.005, ^****^ P< 0.001, one-way ANOVA with Tukey post-hoc test. (**C**) SC samples were obtained by tapes stripping of skin from the inner forearm. Tape strips were mounted on a microscope object slide, while the skin sample was oriented outside. Samples a imaged by AFM in contact mode without further preparation. Overview AFM images of healthy control skin, and skin samples obtained from CPNL and AD patients. Scale bar = 20 µm. (**D**) Quantitative evaluation of the AFM images indicate that the area of corneocytes of lesional AD skin was significantly smaller compared to all other groups. ^**^ P< 0.01, ^***^ P< 0.005, ^****^ P< 0.001, one-way ANOVA with Tukey post-hoc test. (**E**) The corneocyte area correlate significantly with the TEWL. P < 0.001, two-tailed Pearson correlation test. (**F**) Three-dimensional, high resolution AFM images indicate distinct cellular protrusion, previously identified as corneodesmosomes, on corneocytes of AD patients but not healthy controls. (**G**) Quantitative evaluation of AFM images indicate significantly more corneodesmosomes on corneocytes of AD patients. ^**^ P< 0.01, ^***^ P< 0.005, ^****^ P< 0.001, one-way ANOVA with Tukey post-hoc test; ns = not significant.

In AD patients, itch is potentially induced through the penetration of the skin by allergens or other pro-inflammatory compounds due to a reduced skin barrier. To measure the skin barrier, we performed AFM on SC samples obtained from the inner forearm of both patient groups and healthy controls by tape stripping. As schematically illustrated, AFM imaging based on the scanning of the sample by a nanometric probe enabling topographic images of the SC at a nanometric resolution in all three spatial dimensions (Schematic overview, **Figure 2C**). AFM images displayed in **Figure 2C**, indicate that the corneocytes with the SC of healthy and CPNL patient-derived skin are tightly connected and linked through bundles of intermediate filaments (filamentous structures traversing the imaged SC layer). Consistent with a supposed impaired skin barrier, corneocytes were loosely connected in the lesional skin of AD patients and intermediate filaments were absent. Compared to the healthy controls, corneocytes in the lesional skin of AD patients were significantly smaller, whereas no significant changes were measured in the other groups (**Figure 2D**). Cell shrinkage suggests a reduced corneocyte hydration due to an increased water evaporation from skin with an impaired barrier.^19-21^ In line with this, **Figure 2D** shows a clear linear correlation between the trans-epidermal water loss (TEWL) and the corneocyte area. We and other have previously shown that corneocytes of diseased skin are covered by corneodesmosomes^7,22^. Corneodesmosomes are involved in the process of desquamation, which is essential to maintain a fairly constant balance between new and old cells that must be removed^23^. Prior their transition to corneocytes, keratinocytes from the stratum granulosum become densely covered with corneodesmosomes^24^. After transition, corneodesmosomes of healthy SC undergo progressive proteolysis, with the exception of those located at the rims of the flattened corneocytes^25^. In contrast to the healthy SC, we found corneodesmosomes at the corneocyte surface as nanometric protrusions with an average height of 113 ± 30.1 nm and an average width of 695 ± 110.7 nm (n=26) (**Figure 2F**). In line with the corneocyte shrinkage, AD lesional skin showed significantly increased corneodesmosome density (**Figure 2G**). Although the corneodesmosome density was slightly increased in itching skin regions of CPNL patients (CPNL_pruritic_), the difference was not significantly different compared to the healthy controls.

To verify our AFM data, we stained the tape-stripped SC using indirect immunofluorescence with a primary antibody targeting corneodesmosin (CDSN), a key component of corneodesmosomes. **Figure 3A** shows representative images. Consistent with our AFM data, AD lesional skin was characterized by smaller corneocytes and a CDSN staining relocated from the cell-cell contacts to the cellular body. We compared the corneocyte area and corneodemosome density (mean fluorescence intensity per corneocyte) measured by fluorescence microscopy (FLM) and AFM and found a significant correlation of both data sets (**Figure 3B**).

**Figure 3:**
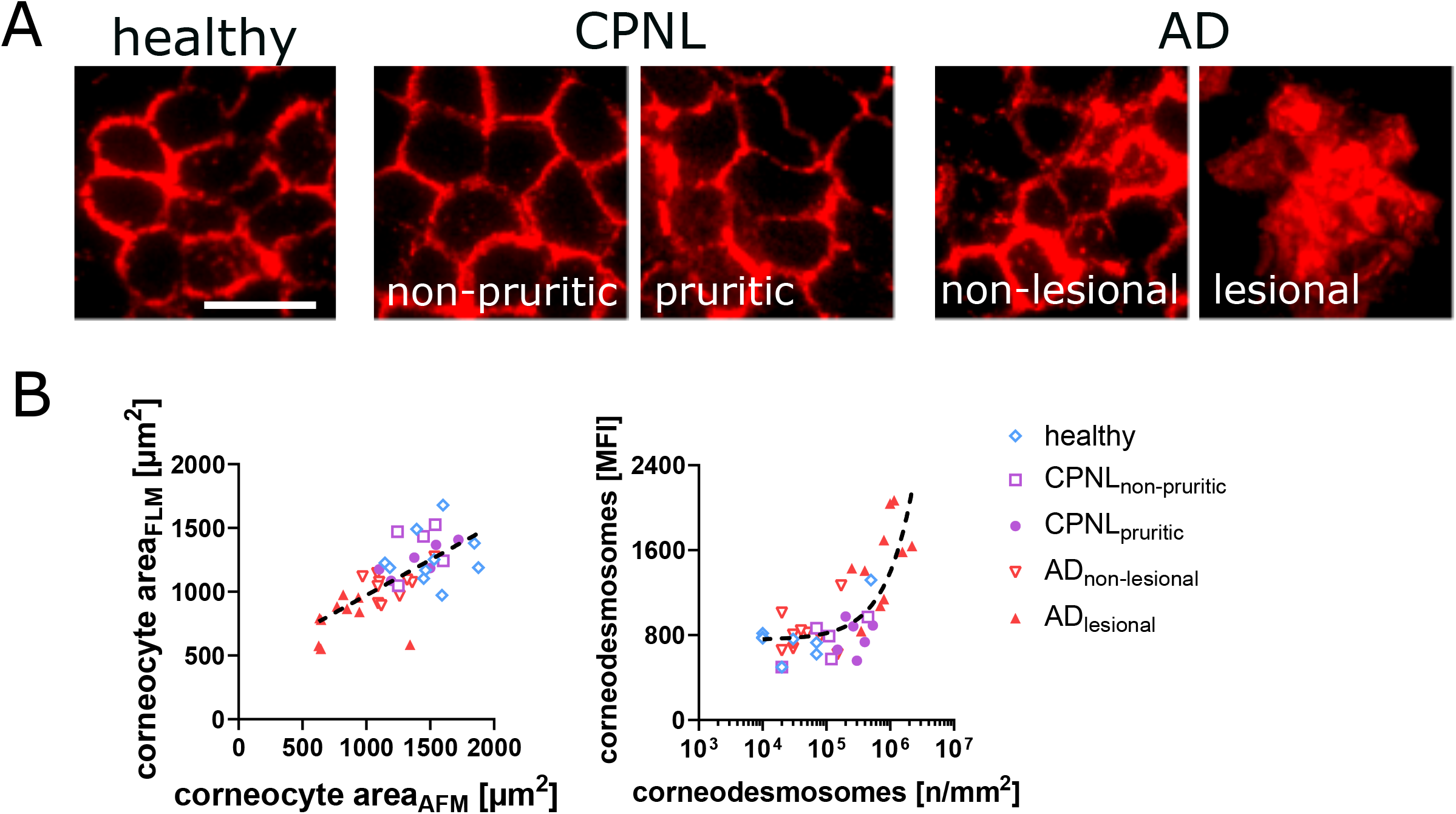
FLM imaging of the SC verified AFM images. (**A**) Representative FLM images of SC samples. CDSN was stained to identify cell-cell contacts of corneocytes and the density (fluorescence intensity) of corneodesmosomes. Scale bar = 50 µm. (**B**) Correlation of the corneocyte areas and the corneodesmosome densities, which have been either measured by AFM or FLM. *P* < 0.001, two-tailed Pearson correlation test.

Finally, we aim to link the morphological changes of the SC with the NRS (**Figure 4A**). Interestingly, by plotting the corneocyte area as a function of the NRS, we found a weak linear positive correlation in the AD and the CPNL patient cohort. This suggest that skin itching, also in AD patients, is not directly connected to the severity of the skin barrier impairment but may follow an all-or-nothing principle. To confirm this, we analyzed AD patients after therapeutic intervention (JAKinhibitors (Abrocitinib), IL-4R antibodies (Dupilumab) or IL-13 antibodies (lebrikizumab)). In these patients, we found that significant therapeutic response was associated with a normalization of the skin barrier (increased corneocyte size) and a strongly reduced itch sensation (**Figure 4B**).

**Figure 4:**
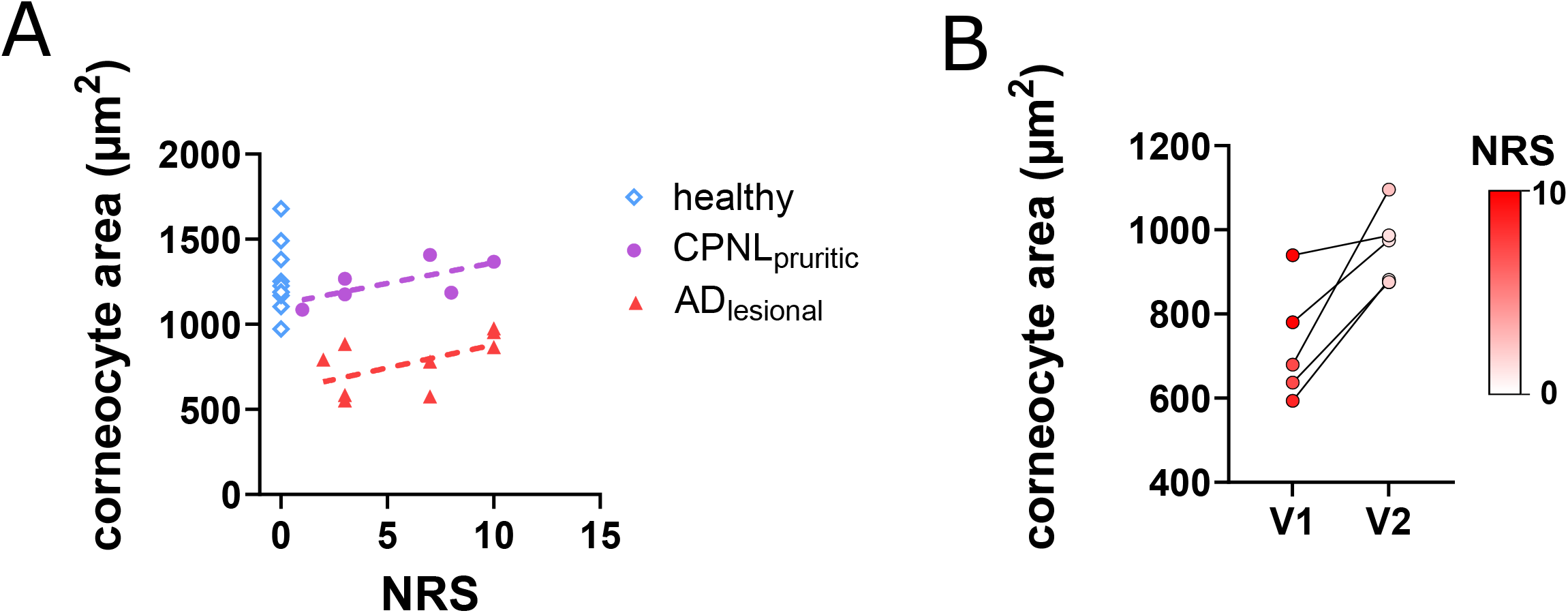
Correlation of the corneocyte area as a function of the NRS. (**A**) Corneocyte areas were different in the distinct groups (healthy, CPNL_pruritic_ and AD_lesional_). Correlations (dashed lines) of the corneocytes within the groups with the NRS indicate no significant differences. (**B**) Treatment of AD patients for 6 month (V1 = visit 1; V2 = visit 2) with JAK inhibitors, IL-4R or IL-13 antibodies restored skin barrier as indicated by the normalization of the corneocyte area (P = 0.012, Student’s t test), and a significantly reduced NRS (P = 0.00001, Student’s t test). The NRS is color coded as indicated by the color gradient bar.

## Discussion

In our study, morphological comparisons between lesional, pruritic and non-lesional, non-pruritic skin in patients with AD revealed significant differences in mean corneocyte area, as well as in the number and localization of corneodesmosomes. Previous research has shown that corneodesmosomes originate from transitional desmosomes, which initially connect corneocytes of the SC to keratinocytes of the granular layer. These corneodesmosomes subsequently link adjacent corneocytes within the SC, contributing to the integrity of the epidermal barrier. During desquamation, corneocytes are shed from the SC, providing both chemical and mechanical protection. This process also supports water impermeability, aided by structural components such as the protein and lipid envelopes of corneocytes.^26^ Three proteins can be found as components of corneodesmosomes: the two desmosomal cadherins, desmoglein 1 (DSG1) and desmocollin 1 (DSC1), and CDSN. ^27^ The stratum conreum continuously undergoes desquamation, a process of inconspicuous shedding of single corneocytes from the skin surface.^28^ It occurs via corneodesmolysis, which stands for proteolysis of the SC corneodesmosomes. ^29^ It has been evidenced in previous studies that progressive proteolysis of CDSN is necessary for the desquamation of corneodesmosomes since it plays a significant role in corneocyte cohesion.^30^ Desquamation of the SC is regulated by a multitude of different proteases, such as kallikreins ^27,31^. Under physiological conditions protease activity is controlled by the extracellular pH. A shift from an acidic to an alkaline pH is observed in lesional atopic skin which leads to increased protease activity and, consequently, enhanced desquamation.^29,32^ Accordingly, corneodesmosomes of healthy cells are only present at the borders of corneocytes, thereby delineating individual cells and enabling accurate quantification of both CDSN intensity and corneocyte size.

Previous research indicated a clear correlation between skin barrier impairment and skin inflammation in AD patients.^33^ Interestingly, although we found clear morphological alterations in the SC of AD patients and an overall high NRS and SSS, we found no correlation between NRS and corneocyte area. This suggests that even minor skin barrier defects may trigger severe pruritus. Interestingly, patients with CPNL reported moderate to high NRS scores despite having SC morphology comparable to healthy skin. This finding further supports the hypothesis that pruritic symptoms can occur independently of barrier disruption.

In conclusion, our results demonstrate that patients with CPNL, despite often exhibiting symptoms similar to those of AD, possess skin morphology nearly identical to that of healthy individuals. A key clinical implication is that our findings do not support the guideline-recommended use of topical emollients for this patient group. Our study provides no evidence of a potential link between CPNL and morphological skin changes, establishing a basis for future research.

## Data availability

All data produced in the present study are available upon reasonable request to the authors

